# The impact of the Black Summer bushfires and other natural disasters on psychological distress of young people: a population-based cohort analysis based on the Longitudinal Study of Australian Children

**DOI:** 10.1101/2025.06.25.25330255

**Authors:** Ben Edwards, Paul Campbell, Matthew Gray

**Author notes:** Corresponding author: Ben Edwards. Funding source: The work was conducted as part of an MHMRC Medical Research Future Fund grant (Grant ID: MRF1201335). Conflict of interest: The authors have no real or perceived conflict of interest to declare.

## Abstract

**Objectives:** To assess the impact of the Australian “Black Summer” bushfires and other disasters of 2019-2020 on adolescent psychological distress. For those exposed to a disaster test the types of disaster impacts associated with worse psychological distress.

**Study design:** Prospective, population-based cohort study; analysis of Longitudinal Study of Australian Children (LSAC) survey data

**Setting, participants:** Adolescents in the nationally representative cross-sequential sample of Australian children recruited in 2004 for the Birth and Kindergarten cohort (aged 0-1 and 4-5 years at enrolment). Survey data from waves 9 (16-17 and 18-19 years for B and K Cohort) and waves 4 to 8 (8-10 and 10-11 years).

**Main outcome measures:** Psychological distress measured by the Kessler 10-item questionnaire, K10.

**Results:** There were 2,726 respondents from the B and K Cohorts who had psychological distress information and demographic characteristics in Wave 9. At wave 9 10% were exposed to bushfires in the 12-months prior, 9% to a storm or cyclone, 18% to at least one disaster and, 7% exposed to more than one disaster. In the previous 10 years 28% had a disaster exposure (waves 4 to 8). The most common disaster impacts were threats to property (40%), impacts on holiday plans (31 %) and advised to evacuate (17%). Exposure to bushfires and storm/cyclone were associated with higher distress in a linear regression model controlling for both demographic characteristics and previous exposure to disasters. For those exposed to a disaster or bushfire those who were advised to evacuate had significantly higher distress in a linear regression model with the same covariates.

**Conclusions:** One-in-ten Australian adolescents were exposed to bushfires in 2019-2020 putting them at risk of a prolonged period of elevated psychological distress. For those who were advised to evacuate distress was even more pronounced and it is important to recognise that further mental health support may still be required.

## Introduction

Lasting 3 months and estimated to have burnt over 24 million hectares,the Black Summer bushfires destroyed over 3,000 buildings and directly killed at least 34 people (Binski, Bennett, & Macintosh, 2020). Around 14.4 per cent of the adult population reported direct exposure to the fires either through their property being damaged or threatened, or being told to evacuate. This equates to around 2.9 million adult Australians (Biddle, Edwards, Herz, & Makkai, 2020).

Much less is known about the mental health impacts of the Black Summer bushfires, studies that have used convenience sample surveys have reported higher depression, anxiety and stress for those affected compared to those with lower levels of exposure (MacLeod et al., 2023) or clinical cut-offs (Usher et al., 2022).

There is even more limited evidence on the impacts of Black Summer on children and adolescents with only one large, but not representative study of adolescents suggesting that those exposed to fires had greater risks for suicidal ideation, trauma and insomnia (Beames, J.R., Huckvale, K., Fujimoto, H. *et al* 2023). Recent longitudinal research in Australia does suggest that fires or floods are associated increased risk of self-harm and suicidal ideation in adolescents 14-19 years however for younger children single disaster exposures such as bushfires were not associated with mental health of children (Edwards, Taylor & Gray, 2024; Campbell, Edwards & Gray, 2024).

Given the scale and scope of the Black Summer bushfires further nationally representative studies are needed. This paper fills this gap in our understanding by examining the impact of the Australian “Black Summer” bushfires of 2019-2020 on adolescent psychological distress. It compares the impact of bushfires on distress while taking into account the impact of other natural disasters and the pandemic. Furthermore, it directly assesses the relationship between distress and a range of common disaster impacts, including property damage and being advised to evacuate.

## Method

### Study design and participants

This study used wave 9 of the Longitudinal Study of Australian Children (LSAC). LSAC, also known as *Growing Up in Australia*, is a large, longitudinal, nationally representative cohort study of Australian children. The survey has been administered every two years since 2004 (wave 1), with wave 9 administered in 2020-2021. The survey has followed two cohorts, the B-Cohort (birth cohort, born in the year preceding wave 1), aged 16-17 in wave 9, and the K-cohort (kindergarten cohort, aged 4-5 in wave 1), aged 20-21 in wave 9 (Growing up in Australia, 2021).

Wave 9 C1 was in field from October-December 2020 (ADA, 2022) and captured exposure to the Australian “Black Summer” bushfires of 2019-20. Waves 1 through 8 consisted of in-person interviews however due to the pandemic the Australian Bureau of Statistics interviewers were not able to conduct face-to-face interviews. Instead, young people and parents were invited to complete an online survey and the fieldwork period was shorter in length than the six-month period. Wave 9C1 consisted of 2,017 respondents from the B-cohort, and 1,786 respondents from the K-cohort.

Disaster exposure from waves 4 to 8 was used as a control variable in early analysis. Results are given in the Appendix.

The original sample of children was drawn using Australia’s public healthcare (Medicare) enrolment database, and the sample is nationally representative of children within each cohort. At the time of sample selection, the Medicare dataset was estimated to cover virtually all four-year old children and 88.5% of children under 12-months (Soloff, Lawrence & Johnstone, 2005). Cross-sectional sample weights, provided by the data custodians to address non-response error, were used in analysis.

The Australian Institute of Family Studies Ethics Committee provided ethics approval for the LSAC, and all participants provided written informed consent.

### Dependent variables

The key dependent variable was psychological distress, measured by the Kessler 10-item questionnaire, K10. Developed by Kessler and others to use in the US National Health Interview Survey measure psychological distress (Kessler, Andrews, Colpe et al., 2002), the K10 has been widely used in screening in population health surveys in Australia (Slade, Grove & Burgess, 2011).

### Disaster exposure variables

The LSAC has collected data on exposure to natural disasters since wave 4. In wave 9, the survey asked adolescents “Have you been affected by any of the following extreme weather events or natural disasters in the past 12 months? Bushfire; Drought; Flood; Storm/Hail; Cyclone; Other extreme weather events or natural disasters”. The survey yields six corresponding self-report measures: one for each type of disaster. In waves 4 to 8 primary caregivers’ reported on whether they have been affected by natural disasters in the previous year. In these waves, caregivers reported on whether in the last 12 months caregivers “had your home or local area affected by bushfire, flooding or a severe storm”, and whether in the past 12 months the caregiver “lived in a drought-affected area”.

### Disaster impact variables

Participants who indicated being exposed to at least one disaster in the past 12 months were asked about the impacts of disaster exposure. Yes/no responses were collected for each of the following:

1. My home or property (including pets or livestock) was damaged or destroyed
2. My home or property was threatened but not damaged or destroyed
3. I was advised by emergency services to evacuate from the area in which I was living or staying
4. My travel plans or my holiday itself were affected
5. My mental and / or physical health was affected

Note that these disaster impact questions were asked of disaster-exposed individuals once only, and for those exposed to more than one disaster, we cannot distinguish which impacts relate to which disasters. There is a many-to-many relationship between disaster exposure and disaster impacts in this dataset.

### Covariates

Statistical models included child and family/neighbourhood characteristics as covariates. Child characteristics were: sex, whether the child was born overseas, whether the child is Aboriginal or Torres Strait Islander, and the cohort to which the child belongs (B-cohort: aged 16-17; or K-cohort: aged 20-21). Family/neighbourhood characteristics included whether the family had moved house since the last wave, whether they lived in a regional or metropolitan area, and their state of residence. Given that policy restrictions and COVID-19 outbreaks varied substantially by states and territories, regional and state of residence dummies also serve to control for variation in experiences in the pandemic (Edwards et al., 2022). Neighbourhood socioeconomic characteristics were measured using Socio- Economic Indexes for Areas (SEIFA) scores (ABS, 2022). We used deciles of the SEIFA Index of Relative Socioeconomic Advantage/Disadvantage, a summary measure of the social and economic conditions of Australian neighborhoods. While we controlled for residential mobility since the last wave there may be many other factors associated with moving to a more disaster-prone area and therefore, we included exposure to disasters in the prior decade as a covariate. This also took account the long-term impacts of disasters so that the mental health impacts of Black Summer and other disaster exposures in the prior 12-month period are identified.

### Statistical analyses

Three sets of linear regression models predicting psychological distress were run. The first assessed the impact of exposure to bushfires (and other natural disasters) on psychological distress. The second analysed those exposed to a disaster, and those exposed to bushfire, to assess the relationship between psychological distress and disaster impacts. All three sets of models controlled for demographic and geographic covariates outlined above.

Logistic regressions were also run on each of the disaster impact variables, in order to understand relationships between disaster type and disaster impact. Note that self-report impacts on mental and/or physical health were deemed to be conceptually too similar to psychological distress to be included in models of distress.

All models were run with the provided cross-sectional sample weights, which correct for survey non-response using Stata 15.

### Ethics approval

The Longitudinal Study of Australian Children was approved by the Australian Institute of Family Studies Ethics Committee.

## Results

In total there were 2,726 respondents who had psychological distress information and demographic characteristics. Table 1 shows rates of exposure to natural disasters, along with impacts reported for those exposed, along with demographic and geographic characteristics of the weighted sample. The table shows that 10.2% of adolescents were exposed to bushfires in the 12-months prior to the survey, and 8.8% were exposed to a storm or cyclone. 18.3% were exposed to at least one disaster, 6.8% exposed to more than one disaster, and 27.7% had previously identified a disaster exposure (in waves 4 to 8). For those exposed to a disaster in wave 9, the most common disaster impact reported was threats to property (40.0%), followed by impacts on holiday plans (31.0%) and physical/mental health (25.9%).

**Table 1:**
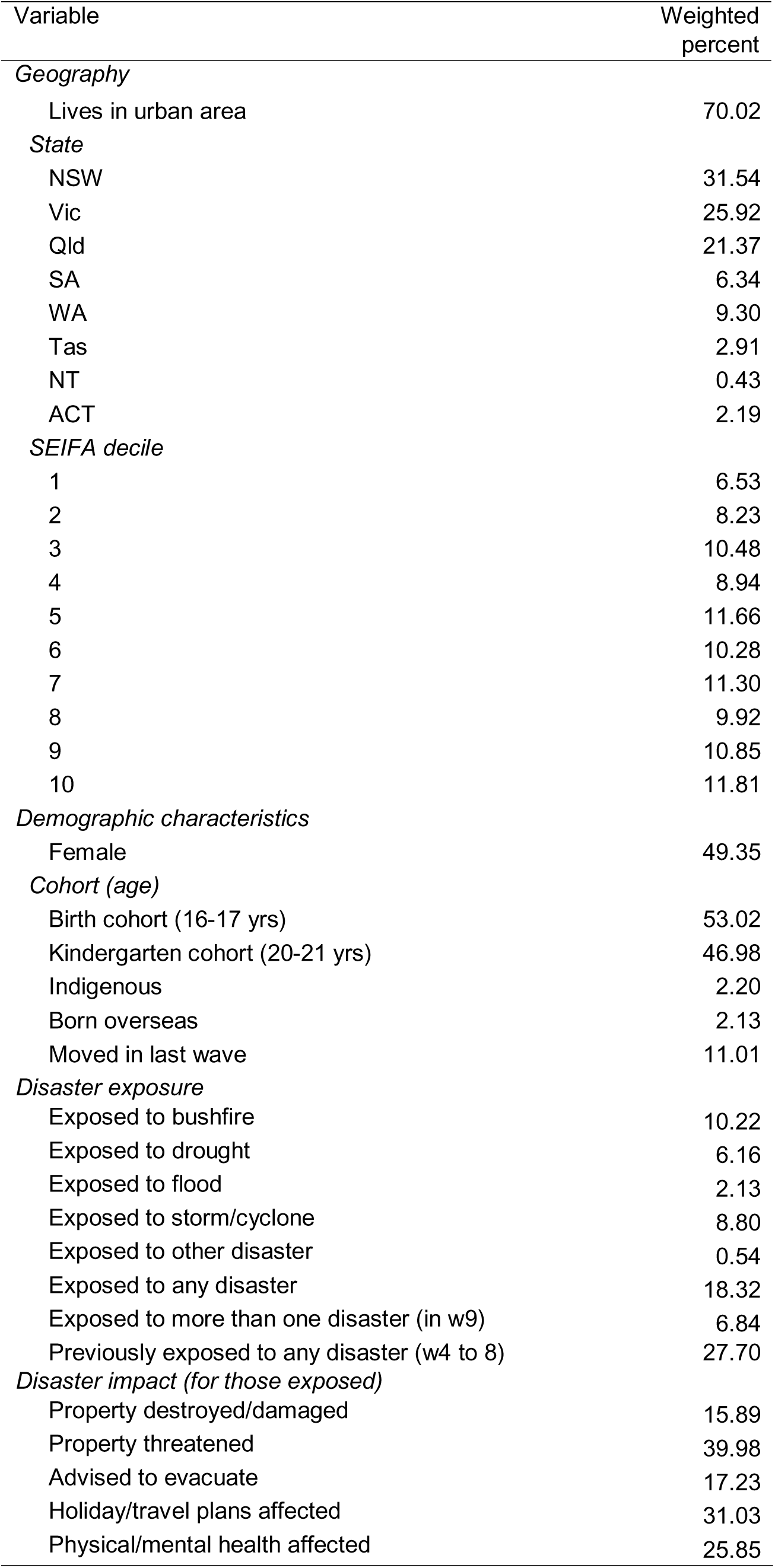
Demographic, geographic characteristics and disaster exposure rates, weighted percentage.

Table 2 shows regression results for disaster exposure predicting psychological distress. Those exposed to a bushfire or a storm/cyclone had significantly higher psychological distress, controlling for both demographic characteristics and previous exposure to disasters (waves 4 to 8).

**Table 2:** Self-report disaster exposure predicting psychological distress (K10), weighted linear regression.

| Variable | Unstd. coeff<br>(B) | Std error | Std. coeff (beta) |
| --- | --- | --- | --- |
| *Disaster exposure* |  |  |  |
| Affected by bushfire | 2.360*** | 0.773 | 0.087 |
| Affected by drought | -0.113 | 1.022 | -0.003 |
| Affected by flood | 1.740 | 1.493 | 0.030 |
| Affected by storm/cyclone | 2.004*** | 0.759 | 0.070 |
| Affected by other disaster | 2.810 | 3.303 | 0.026 |
| Previously exposed to any disaster (w4 to 8) | -0.104 | 0.573 | -0.005 |
| *Demographic controls* |  |  |  |
| Sex (female) | 4.572*** | 0.446 | 0.246 |
| K cohort (aged 20-21) | -0.789* | 0.464 | -0.042 |
| Urban area | 1.057 | 0.659 | 0.052 |
| Indigenous | 0.964 | 1.698 | 0.013 |
| SEIFA decile | -0.138 | 0.090 | -0.041 |
| Moved in past wave | 1.557** | 0.655 | 0.059 |
| Born overseas | 0.379 | 1.276 | 0.006 |
| *State of residence* |  |  |  |
| Vic | 0.546 | 0.576 | 0.026 |
| Qld | 1.079 | 0.688 | 0.048 |
| SA | -0.008 | 0.944 | 0.000 |
| WA | 1.907** | 0.876 | 0.060 |
| Tas | -1.136 | 1.485 | -0.021 |
| NT | 4.484 | 2.800 | 0.032 |
| ACT | -0.487 | 1.096 | -0.008 |
| constant | 19.242*** | 0.916 | . |

N=2,726
\* $p < 0.1$ ; \*\* $p < 0.05$ ; \*\*\* $p < 0.01$
**Base case:** Not affected by natural disaster, male, B-cohort (aged 16-17), lived in regional/remote area, non-Indigenous, did not move in the past wave, born in Australia, resides in NSW.

Table 3 shows relationships between disaster impacts and psychological distress, firstly for those exposed to at least one disaster, and secondly for those exposed to a bushfire. Full regression results (including covariates) can be found in the appendix. Those who were advised to evacuate reported much higher psychological distress. This was the only disaster impact that significantly predicted distress, noting that self-report impacts on physical and mental health was excluded from the models.

**Table 3:** Self-report disaster impacts predicting psychological distress (K10) for those exposed to (a) any disaster and (b) bushfire in the past 12 months, weighted linear regression.

| Variable |  | Unstd. coeff (B) | Std error | Std. coeff (beta) |
| --- | --- | --- | --- | --- |
| *Disaster impacts* |  |  |  |  |
| Any disaster | Property damaged or destroyed | 1.183 | 1.154 | 0.045 |
|  | Property threatened | 0.444 | 0.973 | 0.023 |
|  | Advised to evacuate | 4.295*** | 1.438 | 0.167 |
|  | Holiday plans affected | -0.379 | 1.127 | -0.018 |
| *N=689* |  |  |  |  |
| Bushfire | Property damaged or destroyed | 1.832 | 2.263 | 0.059 |
|  | Property threatened | -0.604 | 1.359 | -0.031 |
|  | Advised to evacuate | 3.672** | 1.567 | 0.175 |
|  | Holiday plans affected | -1.427 | 1.406 | -0.073 |
| *N=383* |  |  |  |  |

\* $p < 0.1$ ; \*\* $p < 0.05$ ; \*\*\* $p < 0.01$
**Base case:** No property damage, property threatened, or holiday plans affected, not advised to evacuate, male, B-cohort (aged 16-17), lived in regional/remote area, non-Indigenous, did not move in the past wave, born in Australia, resides in NSW

There was a strong association between being exposed to bushfire and being advised to evacuate. Among those exposed to a bushfire in the last 12 months, 30.7% were advised to evacuate, and among those advised to evacuate, 98.2% were exposed to a bushfire. The appendix provides details of the relationship between disaster exposure and impact is estimated with regression models, controlling for demographic and geographic characteristics.

## Discussion

In this study we estimated 10% of 16-17 and 20-21 year olds were exposed to the Black Summer bushfires, which is consistent with other estimates. Ten to twelve months later youth were still reporting significantly higher levels of psychological distress than those not exposed to bushfires. Nine percent of youth were also exposed to significant cyclones or storms during this period which also was associated with elevated levels of psychological distress. It is important to highlight that our statistical models took into account prior disaster exposure and other disaster exposure in the 12 months prior as well as demographic characteristics. Variations in pandemic experiences were also accounted for by controlling for state and territory residence and living in regional areas. For those who were exposed to a disaster, there was a significant association between being advised to evacuate and psychological distress, among both the populations exposed to any disaster and those exposed to a bushfire. Given that almost everyone advised to evacuate was exposed to a bushfire, being advised to evacuate nonetheless predicts distress among the subset exposed to a bushfire. Being advised to evacuate may reflect a more severe and imminent threat as reports of property damage was not associated with distress and is consistent with other recent studies suggesting that evacuations though absolutely necessary, as associated with higher levels of psychological distress (ANU work, Biddle).

This is the first national study examining the impacts of the Black Summer bushfires on youth in Australia. Our findings are consistent with the only previous convenience sample of adolescents that reported some adverse mental health impacts (Beames, J.R., Huckvale, K., Fujimoto, H. *et al* 2023) and recent longitudinal research using the same study that reported increased self-harm and suicidal ideation of older adolescents (Edwards, Taylor & Gray, 2024) but no detrimental impacts of single disaster exposures on younger adolescents ( Campbell, Edwards & Gray, 2024).

### Strengths and limitations

This study has several strengths in that it uses a nationally representative sample, collects detailed information about disaster exposures and their impacts and controls for disaster exposures in the previous decade along with other plausible covariates. There are a few limitations in the assessment of disaster exposures however as reports by youth in Wave 9C and by caregivers in the prior decade were limited to the previous 12 months rather than the full two-year period between waves. Consequently, there may well be some disaster exposures that were not captured, leading to some youth who were exposed to a disaster in the 13-to-24-month period in the comparison group, leading to an under-estimation of the full impact of exposures and other natural disasters. Another limitation of the study is that data was being collected during the midst of a pandemic, but despite the huge variation in policy responses and outbreaks by state and territories (Edwards, et al., 2022) there were few significant differences by state and territory and regional indicators. One other consequence of the pandemic was the increased propensity for young people to move back with parents (AIFS research) which was captured in our statistical modelling and associated with higher levels of psychological distress.

## Conclusion

In this study we estimate that one-in-ten Australian youth were exposed to the Black Summer bushfires and that this exposure had detrimental mental health impacts ten to twelve months later. Those who were considered by authorities to be sufficiently at risk to warrant being warned to evacuate were worst effected, reflecting the level of exposure to imminent harm. Future research needs to understand whether this particular fire had significant longer-term impacts such as the Black Saturday bushfires in Victoria (Bryant, et al. 2014) and to identify potential protective factors that policies can influence that mitigate the mental health risks.

## Supporting information

STROBE Checklist Cohort

## Data Availability

Data are available through the Australian Data Archives, details are below:
Department of Social Services; Australian Institute of Family Studies; Australian Bureau of Statistics, 2022, "Growing Up in Australia: Longitudinal Study of Australian Children (LSAC) Release 9.1 C2 (Waves 1-9C)", https://doi.org/10.26193/QR4L6Q, ADA Dataverse, V5

## Appendix

Full regression model for Table 3 (part 1): Self-report disaster impacts predicting psychological distress (K10) for those exposed to any disaster in the past 12 months, weighted linear regression

| Variable | Unstd. coeff (B) | Std error | Std. coeff (beta) |
| --- | --- | --- | --- |
| *Disaster impacts* |  |  |  |
| Property damaged or destroyed | 1.183 | 1.154 | 0.045 |
| Property threatened | 0.444 | 0.973 | 0.023 |
| Advised to evacuate | 4.295*** | 1.438 | 0.167 |
| Holiday plans affected | -0.379 | 1.127 | -0.018 |
| *Demographic controls* |  |  |  |
| Sex (female) | 3.989*** | 0.945 | 0.205 |
| K cohort (aged 20-21) | -2.088** | 1.018 | -0.106 |
| Urban area | 3.527*** | 1.192 | 0.180 |
| Indigenous | -5.330*** | 1.985 | -0.042 |
| SEIFA decile | -0.478** | 0.217 | -0.135 |
| Moved in past wave | 2.547* | 1.418 | 0.091 |
| Born overseas | -3.715 | 2.288 | -0.047 |
| *State of residence* |  |  |  |
| Vic | 0.577 | 1.267 | 0.022 |
| Qld | 2.559* | 1.347 | 0.109 |
| SA | 2.194 | 1.885 | 0.044 |
| WA | 2.047 | 2.342 | 0.053 |
| Tas | 1.450 | 2.943 | 0.020 |
| NT | 1.926 | 5.440 | 0.010 |
| ACT | 1.059 | 1.413 | 0.024 |
| constant | 21.668*** | 1.516 | . |

N=689
\* $p < 0.1$ ; \*\* $p < 0.05$ ; \*\*\* $p < 0.01$
**Base case:** No property damage, property threatened, or holiday plans affected, not advised to evacuate, male, B-cohort (aged 16-17), lived in regional/remote area, non-Indigenous, did not move in the past wave, born in Australia, resides in NSW

Full regression model for Table 3 (part 2): Self-report disaster impacts predicting psychological distress (K10) for those exposed to a bushfire in the past 12 months, weighted linear regression

| Variable | Unstd. coeff (B) | Std error | Std. coeff (beta) |
| --- | --- | --- | --- |
| *Disaster impacts* |  |  |  |
| Property damaged or destroyed | 1.832 | 2.263 | 0.059 |
| Property threatened | -0.604 | 1.359 | -0.031 |
| Advised to evacuate | 3.672** | 1.567 | 0.175 |
| Holiday plans affected | -1.427 | 1.406 | -0.073 |
| *Demographic controls* |  |  |  |
| Sex (female) | 4.117*** | 1.368 | 0.210 |
| K cohort (aged 20-21) | -2.372* | 1.256 | -0.121 |
| Urban area | 2.227 | 1.567 | 0.114 |
| Indigenous | -0.731 | 2.164 | -0.006 |
| SEIFA decile | -0.217 | 0.237 | -0.062 |
| Moved in past wave | 2.218 | 1.896 | 0.084 |
| Born overseas | -3.031 | 4.618 | -0.028 |
| *State of residence* |  |  |  |
| Vic | 0.201 | 1.879 | 0.008 |
| Qld | 1.416 | 2.617 | 0.048 |
| SA | 3.028 | 2.144 | 0.065 |
| WA | 5.941 | 3.838 | 0.146 |
| Tas | 3.908 | 3.043 | 0.049 |
| NT | 1.203 | 6.962 | 0.008 |
| ACT | -0.478 | 1.514 | -0.012 |
| constant | 22.217*** | 2.214 | . |

N=383
\* $p < 0.1$ ; \*\* $p < 0.05$ ; \*\*\* $p < 0.01$
**Base case:** No property damage, property threatened, or holiday plans affected, not advised to evacuate, B-cohort (aged 16-17), lived in regional/remote area, non-Indigenous, did not move in the past wave, born in Australia, resides in NSW

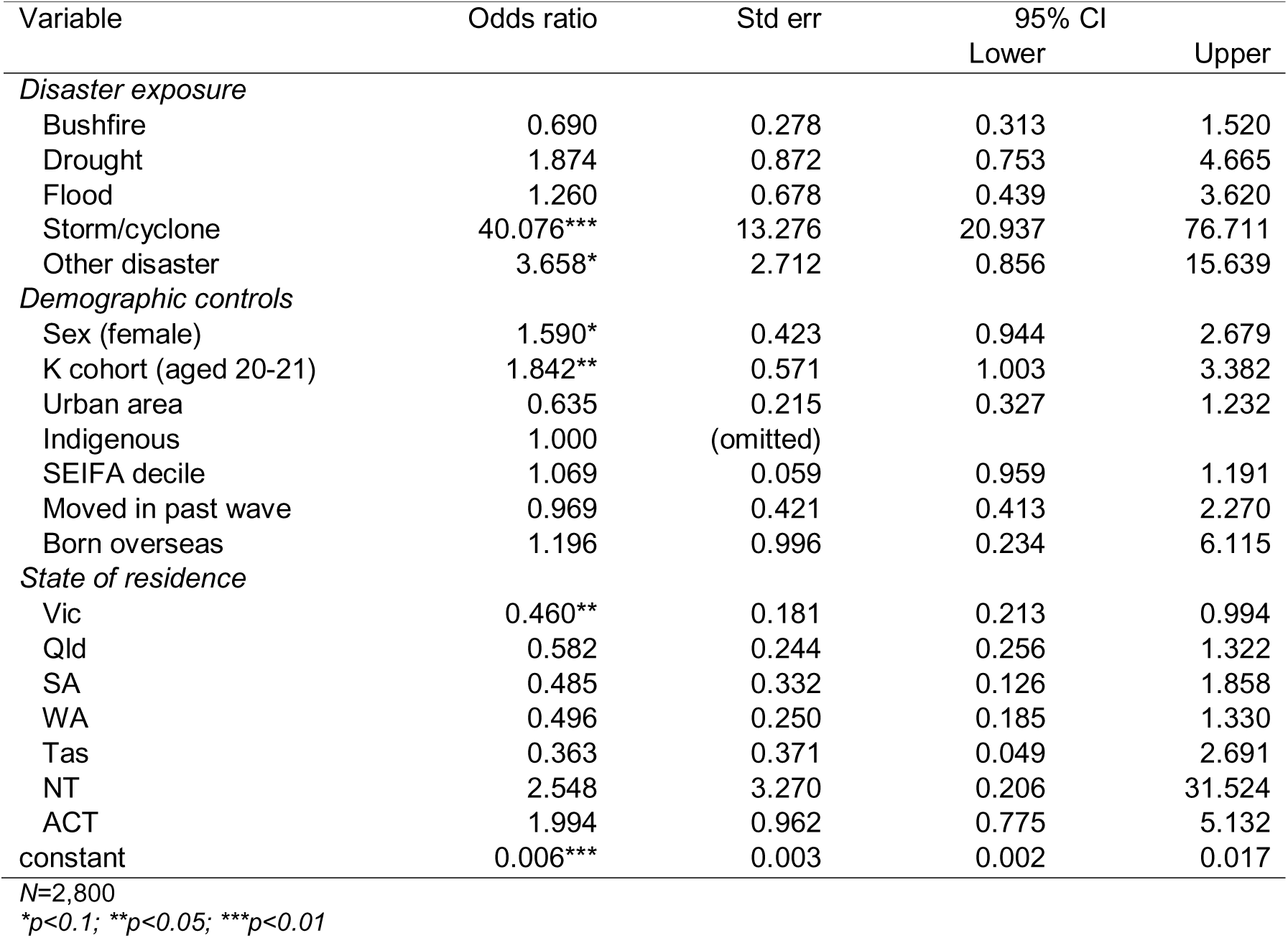
Odds of experiencing property damage

Odds of experiencing threats to property

| Variable | Odds ratio | Std err | 95% CI |  |
| --- | --- | --- | --- | --- |
|  |  |  | Lower | Upper |
| *Disaster exposure* |  |  |  |  |
| Bushfire | 11.556*** | 2.843 | 7.135 | 18.716 |
| Drought | 3.285*** | 1.086 | 1.719 | 6.278 |
| Flood | 1.925 | 1.230 | 0.550 | 6.735 |
| Storm/cyclone | 6.794*** | 1.907 | 3.919 | 11.779 |
| Other disaster | 0.504 | 0.664 | 0.038 | 6.666 |
| *Demographic controls* |  |  |  |  |
| Sex (female) | 0.956 | 0.205 | 0.627 | 1.456 |
| K cohort (aged 20-21) | 1.537* | 0.348 | 0.985 | 2.396 |
| Urban area | 0.306*** | 0.080 | 0.184 | 0.511 |
| Indigenous | 1.674 | 0.996 | 0.521 | 5.376 |
| SEIFA decile | 1.070 | 0.049 | 0.978 | 1.170 |
| Moved in past wave | 0.804 | 0.211 | 0.480 | 1.345 |
| Born overseas | 0.341 | 0.312 | 0.057 | 2.047 |

| *State of residence* |  |  |  |  |
| --- | --- | --- | --- | --- |
| Vic | 0.658 | 0.203 | 0.360 | 1.204 |
| Qld | 1.408 | 0.422 | 0.783 | 2.532 |
| SA | 1.578 | 0.646 | 0.707 | 3.522 |
| WA | 1.831* | 0.574 | 0.990 | 3.386 |
| Tas | 0.617 | 0.262 | 0.268 | 1.419 |
| NT | 0.351 | 0.372 | 0.044 | 2.800 |
| ACT | 1.488 | 0.629 | 0.650 | 3.408 |
| constant | 0.028*** | 0.010 | 0.014 | 0.057 |

*N*=2,832
\**p*<0.1; \*\**p*<0.05; \*\*\**p*<0.01

Odds of experiencing being advised to evacuate

| *Odds of experiencing being advised to evacuate* |  |  |  |  |
| --- | --- | --- | --- | --- |
| Variable | Odds ratio | Std err | 95% CI |  |
|  |  |  | Lower | Upper |
| *Disaster exposure* |  |  |  |  |
| Bushfire | 1512.028*** | 1120.228 | 353.935 | 6459.459 |
| Drought | 1.139 | 0.421 | 0.552 | 2.352 |
| Flood | 1.452 | 0.843 | 0.465 | 4.532 |
| Storm/cyclone | 0.811 | 0.295 | 0.397 | 1.654 |
| Other disaster | 1.113 | 1.207 | 0.133 | 9.328 |
| *Demographic controls* |  |  |  |  |
| Sex (female) | 1.045 | 0.301 | 0.595 | 1.838 |
| K cohort (aged 20-21) | 1.503 | 0.460 | 0.825 | 2.737 |
| Urban area | 0.353*** | 0.119 | 0.183 | 0.683 |
| Indigenous | 1.000 | (omitted) |  |  |
| SEIFA decile | 1.020 | 0.067 | 0.897 | 1.161 |
| Moved in past wave | 0.341** | 0.157 | 0.138 | 0.842 |
| Born overseas | 4.188 | 7.094 | 0.151 | 115.865 |
| *State of residence* |  |  |  |  |
| Vic | 0.830 | 0.349 | 0.365 | 1.891 |
| Qld | 0.398* | 0.218 | 0.136 | 1.166 |
| SA | 2.003 | 1.363 | 0.528 | 7.598 |
| WA | 4.709*** | 2.481 | 1.677 | 13.225 |
| Tas | 0.811 | 0.679 | 0.157 | 4.182 |
| NT | 1.000 | (empty) |  |  |
| ACT | 0.954 | 0.546 | 0.310 | 2.929 |
| constant | 0.000*** | 0.000 | 0.0001 | 0.002 |

*N*=2,780
\**p*<0.1; \*\**p*<0.05; \*\*\**p*<0.01

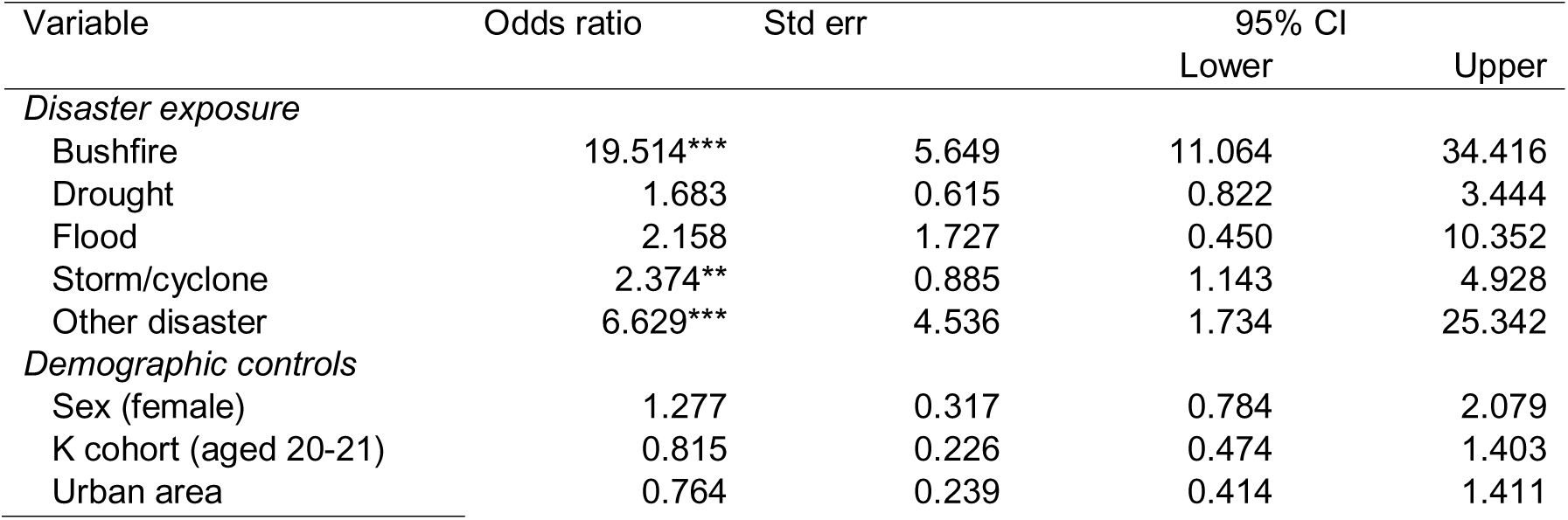
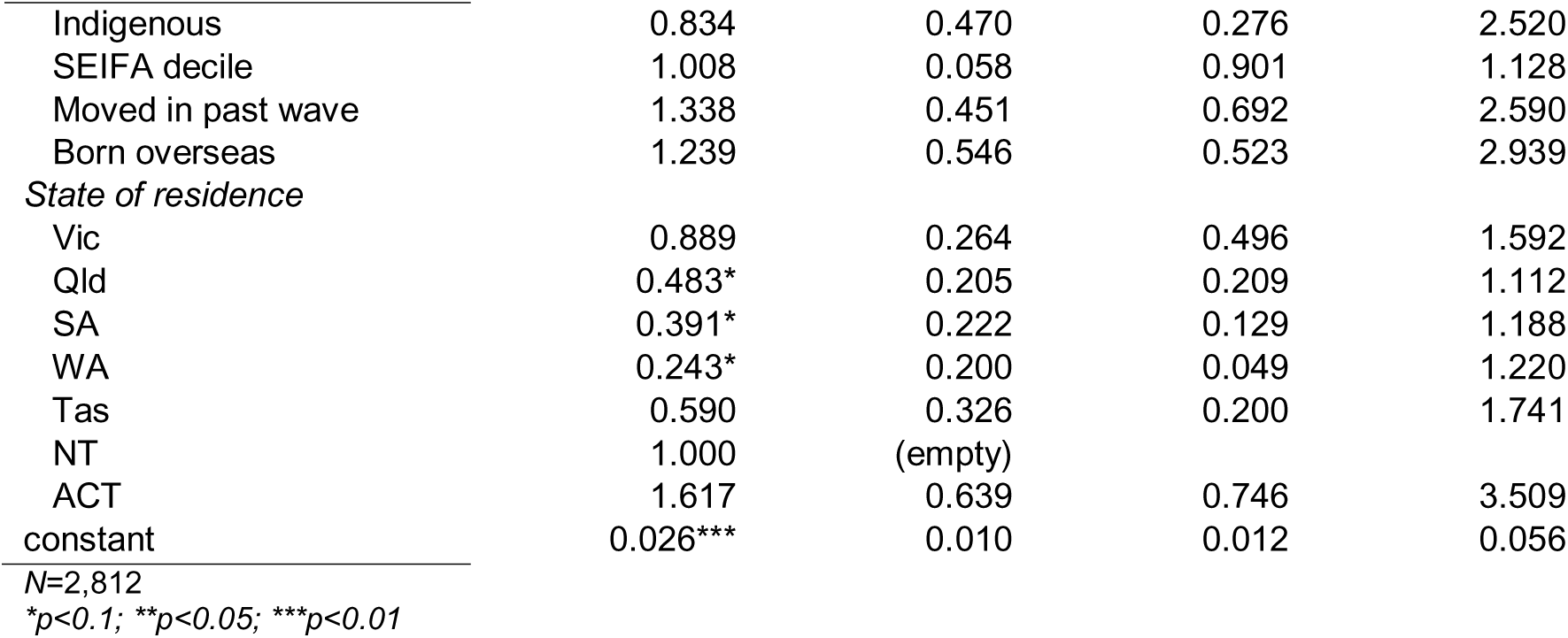
Odds of experiences holiday/travel plans affected

Odds of experiencing impacts on physical or mental health

| Variable | Odds ratio | Std err | 95% CI |  |
| --- | --- | --- | --- | --- |
|  |  |  | Lower | Upper |
| *Disaster exposure* |  |  |  |  |
| Bushfire | 35.928*** | 11.615 | 19.066 | 67.704 |
| Drought | 2.293* | 0.985 | 0.988 | 5.322 |
| Flood | 0.459 | 0.357 | 0.100 | 2.113 |
| Storm/cyclone | 1.486 | 0.501 | 0.767 | 2.879 |
| Other disaster | 9.342* | 12.064 | 0.743 | 117.387 |
| *Demographic controls* |  |  |  |  |
| Sex (female) | 2.158*** | 0.596 | 1.257 | 3.708 |
| K cohort (aged 20-21) | 0.833 | 0.247 | 0.466 | 1.490 |
| Urban area | 0.976 | 0.322 | 0.512 | 1.863 |
| Indigenous | 0.880 | 0.559 | 0.253 | 3.059 |
| SEIFA decile | 0.992 | 0.055 | 0.891 | 1.105 |
| Moved in past wave | 1.660 | 0.551 | 0.866 | 3.182 |
| Born overseas | 2.433 | 1.971 | 0.498 | 11.900 |
| *State of residence* |  |  |  |  |
| Vic | 0.924 | 0.297 | 0.493 | 1.735 |
| Qld | 0.970 | 0.445 | 0.395 | 2.382 |
| SA | 0.476 | 0.217 | 0.195 | 1.165 |
| WA | 0.492 | 0.338 | 0.128 | 1.891 |
| Tas | 0.731 | 0.488 | 0.198 | 2.706 |
| NT | 1.000 | (empty) |  |  |
| ACT | 1.915* | 0.748 | 0.891 | 4.116 |
| constant | 0.008*** | 0.004 | 0.003 | 0.020 |

*N*=2,812
\**p*<0.1; \*\**p*<0.05; \*\*\**p*<0.01

